# A deep learning based graph-transformer for whole slide image classification

**DOI:** 10.1101/2021.10.15.21265060

**Authors:** Yi Zheng, Rushin Gindra, Margrit Betke, Jennifer E. Beane, Vijaya B. Kolachalama

**Affiliations:** Department of Computer Science, College of Arts and Sciences, Boston University, Boston, MA, USA; Department of Medicine, Boston University School of Medicine, Boston, MA, USA

**Keywords:** Digital pathology, Graph convolutional network, Lung cancer, Transformer

## Abstract

Deep learning is a powerful tool for assessing pathology data obtained from digitized biopsy slides. In the context of supervised learning, most methods typically divide a whole slide image (WSI) into patches, aggregate convolutional neural network outcomes on them and estimate overall disease grade. However, patch-based methods introduce label noise in training by assuming that each patch is independent with the same label as the WSI and neglect the important contextual information that is significant in disease grading. Here we present a Graph-Transformer (GT) based framework for processing pathology data, called GTP, that interprets morphological and spatial information at the WSI-level to predict disease grade. To demonstrate the applicability of our approach, we selected 3,024 hematoxylin and eosin WSIs of lung tumors and with normal histology from the Clinical Proteomic Tumor Analysis Consortium, the National Lung Screening Trial, and The Cancer Genome Atlas, and used GTP to distinguish adenocarcinoma (LUAD) and squamous cell carcinoma (LSCC) from those that have normal histology. Our model achieved consistently high performance on binary (tumor versus normal: mean overall accuracy = 0.975 ± 0.013) as well as three-label (normal versus LUAD versus LSCC: mean accuracy = 0.932 ± 0.019) classification on held-out test data, underscoring the power of GT-based deep learning for WSI-level classification. We also introduced a graphbased saliency mapping technique, called GraphCAM, that captures regional as well as contextual information and allows our model to highlight WSI regions that are highly associated with the class label. Taken together, our findings demonstrate GTP as a novel interpretable and effective deep learning framework for WSI-level classification.

## I. Introduction

**C**OMPUTATIONAL pathology [1]–[4], which entails the analysis of digitized biopsies of a bodily tissue, is gaining increased attention over the past few years. The sheer amount of information on a single whole slide image (WSI) typically can exceed over a gigabyte, so traditional image analysis routines may not be able to fully process all this data in an efficient fashion. Modern machine learning methods such as deep learning have allowed us to make great progress in terms of analyzing WSIs including disease classification [5], tissue segmentation [6], mutation prediction [7], spatial profiling of immune infiltration [8], and so on. Most of these methods rely on systematic breakdown of WSIs into image patches, followed by development of deep neural networks at patchlevel and integration of outcomes on these patches to create overall WSI-level estimates. While patch-based approaches catalyzed research in the field, the community has begun to appreciate the conditions in which they confer benefit and in those where they cannot fully capture the underlying pathology. For example, methods focused on identifying the presence or absence of a tumor on an WSI can be developed on patches using computationally efficient techniques such as multiple instance learning [9]. On the other hand, if the goal is to identify the entire tumor region or capture the connectivity of the tumor microenvironment characterizing the stage of disease, then it becomes important to assess both local and regional information on the WSI. There are several other scenarios where both the patch- and WSI-level features need to be identified to assess the pathology [10], and computational methods to perform such analysis are much needed.

The success of patch-based deep learning methods can be attributed to the availability of pre-trained deep neural networks on natural images from public databases (i.e., ImageNet [11]). Since there are millions of parameters in a typical deep neural network, *de novo* training of this network requires access to a large set of pathology data, and such resources are not necessarily available at all locations. To address this bottleneck, researchers have leveraged transfer learning approaches that are pre-trained on ImageNet to accomplish various tasks. Recently, transformer architectures were applied directly to sequences of image patches for various classification tasks. Specifically, Vision Transformers (ViT) were shown to achieve excellent results compared to state-of-the-art convolutional networks while requiring substantially fewer computational resources for training [12]. Position embeddings were used in ViTs to retain spatial information and capture the association of different patches within the input image. Excitingly, the self-attention mechanism in ViT requires the calculation of pairwise similarity scores on all the patches, resulting in memory efficiency and a simple time complexity that is quadratic in the number of patches. Leveraging such approaches to perform pathology image analysis is not trivial because each WSI can contain thousands of patches. Additionally, some approximations are often made on these patches such as using the WSI-level label on each patch during training, which are not ideal in all scenarios as there is a need to process both the local information as well as the WSI in its entirety to better understand the pathological correlates of disease. Similar to the local and WSI-level examination, we argue that an expert pathologist’s workflow also involves examination of the entire biopsy slide using manual operations such as panning and zooming in and out of specific regions of interest to assess various aspects of disease at multiple scales. In the zoom-in assessment, pathologists perform indepth, microscopic evaluation of local pathology whereas, the zoom-out assessment involves obtaining a rational estimate of the contextual features on the entire WSI. Both these assessments are critical as the pathologist obtains a gestalt on various features to comprehensively assess the disease [10].

Recent attempts to perform WSI-level analysis have shown promising results in terms of assessing the overall tissue microenvironment. In particular, graph-based approaches have gained a lot of traction due to their ability to represent the entire WSI and analyze patterns to predict various outcomes of interest. Zhou and colleagues developed a cell-graph convolutional neural network on WSIs to predict the grade of colorectal cancer (CRC) [13]. In this work, the WSI was converted to a graph, where each nucleus was represented by a node and the cellular interactions were denoted as edges between these nodes to accurately predict CRC grade. Also, Adnan and colleagues developed a two-stage framework for WSI representation learning [14], where patches were sampled based on color and a graph neural network was constructed to learn the inter-patch relationships to discriminate lung adenocarcinoma (LUAD) from lung squamous cell carcinoma (LSCC). In another recent work, Lu and team developed a graph representation of the cellular architecture on the entire WSI to predict the status of human epidermal growth factor receptor 2 and progesterone receptor [15]. Their architecture attempted to create a bottom-up approach (i.e., nuclei- to WSI-level) to construct the graph, and in so doing, achieved a relatively efficient framework for analyzing the entire WSI.

We contend that integration of computationally efficient approaches such as ViTs along with graphs can lead to more efficient approaches for the assessment of WSIs. To address this aspect, we developed a graph-based vision transformer called GTP that leverages the graph-based representation of pathology images and the computational efficiency of transformer architectures to perform WSI-level analysis. The GTP framework involves construction of a graph convolutional network by embedding image patches in feature vectors using contrastive learning, followed by the application of a transformer to predict a WSI-level label corresponding to a specific disease type. We used WSIs from three publicly available data resources to develop a GTP model to distinguish normal WSIs from those with lung tumors. Additionally, we extended our framework to classify normal WSIs from those with LUAD or LSCC. We also introduce graph-based class activation mapping (GraphCAM), a novel approach to generate WSI-level saliency maps that are able to identify image regions that are highly associated with the class label.

## II. Materials and methods

### A. Study population

We obtained access to WSI data of lung tumors (LUAD and LSCC) and normal tissue from the Clinical Proteomic Tumor Analysis Consortium (CPTAC), the National Lung Screening Trial (NLST) and The Cancer Genome Atlas (TCGA) (Table I). CPTAC is a national effort to accelerate the understanding of the molecular basis of cancer through the application of large-scale proteome and genome analysis [16]. NLST was a randomized controlled trial to determine whether screening for lung cancer with low-dose helical computed tomography reduces mortality from lung cancer in high-risk individuals relative to screening with chest radiography [17]. TCGA is a landmark cancer genomics program, which molecularly characterized thousands of primary cancer and matched normal samples spanning 33 cancer types [18]. For each of these cases, we also obtained relevant demographic and clinical information.

**TABLE I:** Study population. Whole slide images and corresponding clinical information from three distinct cohorts including the Clinical Proteomic Tumor Analysis Consortium (CPTAC), The Cancer Genome Atlas (TCGA) and the National Lung Screening Trial (NLST) were used.

| (a) CPTAC |  |
| --- | --- |
| Description | Value |
| Number of patients | 435 |
| Number of whole slide images | 2071 |
| Number of whole slide images per class<sup>1</sup> | 719, 667, 685 |
| Number of patches<sup>2</sup> | 1277, [100-8478] |
| Age<sup>3</sup> | 1, 4, 23, 80, 134, 89, 5, 99 |
| Gender<sup>4</sup> | 235, 101, 99 |
| Race<sup>5</sup> | 89, 5, 1, 1, 339 |
| (b) TCGA |  |
| Description | Value |
| Number of patients | 256 |
| Number of whole slide images | 288 |
| Number of whole slide images per class<sup>1</sup> | 92, 97, 99 |
| Number of patches<sup>2</sup> | 571.5, [100-7570] |
| Age<sup>3</sup> | 10, 1, 12, 42, 93, 84, 16, 8 |
| Gender<sup>4</sup> | 144, 112, 0 |
| Race<sup>5</sup> | 188, 28, 3, 0, 37 |
| (c) NLST |  |
| Description | Value |
| Number of patients | 345 |
| Number of whole slide images | 665 |
| Number of whole slide images per class<sup>1</sup> | 75, 378, 212 |
| Number of patches<sup>2</sup> | 2679.5, [110-7029] |
| Age<sup>3</sup> | 0, 0, 0, 87, 201, 57, 0, 0 |
| Gender<sup>4</sup> | 211, 134, 0 |
| Race<sup>5</sup> | 315, 14, 11, 1, 4 |

<sup>1</sup> Normal, LUAD, LSCC <sup>2</sup> Median, Range
<sup>3</sup> Binned: 20-29, 30-39, 40-49, 50-59, 60-69, 70-79, 80-89, Unknown
<sup>4</sup> Males, Females, Unknown
<sup>5</sup> White, Black or African-American, Asian, American Indian or Alaskan Native, Other/unknown

### B. Graph-Transformer

Our proposed Graph-Transformer (GT) network fuses a graph representation *G* of an WSI and a transformer that can generate WSI-level predictions in a computationally efficient fashion (Figure 1). Let *G* = (*V, E*) be an undirected graph where *V* is the set of nodes representing the image patches and *E* is the set of edges between the nodes in *V* that represent whether two image patches are adjacent to each other. We denote the adjacency matrix of *G* as 𝒜 = [𝒜_*ij*_] where 𝒜_*ij*_ = 1 if there exists an edge (*v*_*i*_, *v*_*j*_) ∈ *E* and 𝒜_*ij*_ = 0 otherwise. An image patch must be connected to other patches and can be surrounded by at most 8 adjacent patches, so the sum of each row or column of is at least one and at most 8. A graph can be associated with a node feature matrix *F*, *F* ∈ IR^*N×D*^, where each row contains the *D*-dimensional feature vector computed for an image patch, i.e. node, and *N* = |*V*|. An example of a WSI, its patches, associated graph, and node feature matrix are illustrated in Figure S1.

**Fig. 1:**
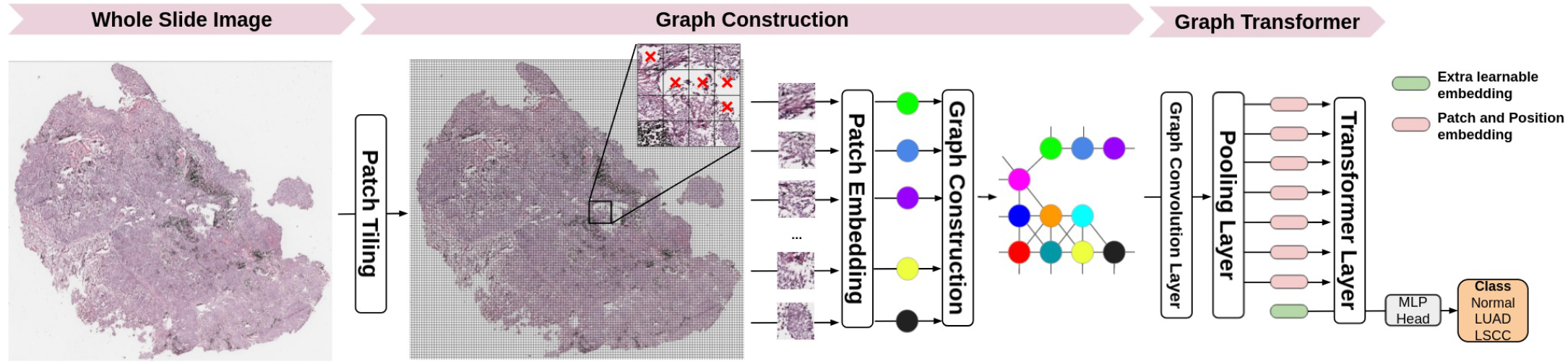
Schematic of GTP deep learning framework. Each whole slide image (WSI) was divided into patches followed by elimination of the patches that predominantly contained the background. Each image patch was then embedded in feature vectors by a contrastive learning-based patch embedding module. The feature vectors were then used to build the graph followed by a transformer that takes the graph as the input and predicts WSI-level class label.

As shown in Fig. 1, given a WSI, the classification task contains two steps, graph construction and graph interpretation by a transformer. The second step aims to learn a mapping from the WSI-associated graph and its node feature matrix to the corresponding label of the WSI.

Using all the pixels within each image patch as features can make model training computationally intractable. Instead, our framework applies a feature extractor to generate a vector containing features and uses it to define the information contained in an image patch, which is a node in the graph. This step reduces the node feature dimension from *W*_*p*_×*H*_*p*_ ×*C*_*p*_ to *D*, where *W*_*p*_, *H*_*p*_, and *C*_*p*_ are width, height, and channel of the image patch, and *D*×1 is the dimension of extracted feature vector. The expectation is that the derived feature vector provides an efficient representation of the node and also serves as a robust means by which to define a uniform representation of an image patch for graph-based classification.

As described above, current methods that have been developed at patch-level impose WSI-level labels on all the patches or use weakly supervised learning to extract feature vectors that are representative of the WSI. This strategy is not suitable for all scenarios, especially when learning the contextual information on the WSI is needed. We leveraged a strategy based on self-supervised contrastive learning [19], to extract features from the WSIs. This framework enables robust representations that can be learned without the need for manual labels. Our approach involves using contrastive learning to train a CNN that produces embedding representations by maximizing agreement between two differently augmented views of the same image patch via a contrastive loss in the latent space (Figure S2). GTP tiles the WSIs from the training set into patches and randomly samples a mini-batch of *K* patches. Two different data augmentation operations are applied to each patch (*p*), resulting i006E two augmented patches (*p*_*i*_ and *p*_*j*_). The pair of two augmented patches from the same patch is denoted as a positive pair. For a mini-batch of *K* patches, there are 2*K* augmented patches in total. Given a positive pair, the other 2*K −* 1 augmented patches are considered as negative samples. Subsequently, our GTP approach uses a CNN to extract representative embedding vectors (*f*_*i*_, *f*_*j*_) from each augmented patch (*p*_*i*_, *p*_*j*_). The embedding vectors are then mapped by a projection head to a latent space (*z*_*i*_, *z*_*j*_) where contrastive leaning loss is applied. The contrastive learning loss function for a positive pair of augmented patches (i, j) is defined as:

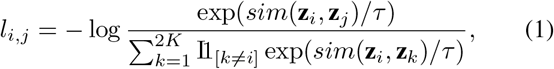

where I1_[*k ≠ i*]_ *∈* {0, 1} is an indicator function evaluating to 1 if and only if *k ≠ i* and *τ* denotes a temperature parameter. Also, *sim*(**u, v**) = **u**^*T*^ **v**/∥**u**∥ ∥**v**∥denotes the dot product between *L*_2_ normalized **u** and **v** (i.e., cosine similarity). For model training, the patches were densely cropped without overlap and treated as individual images. The final loss was computed across all positive pairs, including both (i, j) and (j, i) in a mini-batch. After convergence, we kept the feature extractor and used it for our GTP model to compute the feature vectors of the patches from the WSIs. GTP uses these computed feature vectors as node features in the graph construction phase. Specifically, we obtained the node-specific feature matrix *F* = [*f*_1_; *f*_2_; … ; *f*_*N*_ ], *F* ∈ IR^*N×D*^, where *f*_*i*_ is the D-dimensional embedding vector obtained from Resnet trained using contrastive learning and *N* is the number of patches from one WSI. Note that *N* is variable since different WSIs contain different numbers of patches. As a result, each node in *F* corresponds to one patch of the WSI. We defined an edge between a pair of nodes in *F* based on the spatial location of its corresponding patches on the WSI. If patch *i* is a neighbor of patch *j* on the WSI (Figure S1), then GTP creates an edge between node *i* and node *j* as well as set 𝒜_*ij*_ = 1 and 𝒜_*ji*_ = 1, otherwise 𝒜 _*ij*_ = 0 and 𝒜_*ji*_ = 0. GTP uses feature node matrix *F* and adjacent matrix 𝒜 to construct a graph to represent each WSI.

The Graph Transformer component of GTP consists of a graph convolutional (GC) layer, a transformer layer, and a pooling layer. We implemented the GC layer, introduced by Kipf & Welling [20], to handle the graph-structured data. The GC layer operates message propagation and aggregation in the graph, and is defined as:

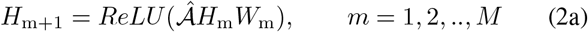

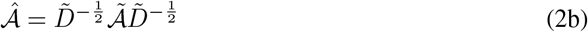

where 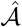 is the symmetric normalized adjacency matrix of 𝒜 and *M* is the number of GC layers. Here, 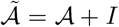 is the adjacency matrix with a self-loop added to each node, and 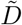 is a diagonal matrix where 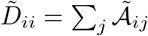. *H*_*m*_ is the input of the *m*-th GC layer and *H*_1_ is initialized with the node feature matrix *F*. Additionally, 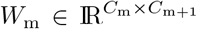 is the matrix of learnable filters in the GC layer, where *C*_m_ is the dimension of the input and *C*_m+1_ is the dimension of the output.

The GC layer of GTP enables learning of node embeddings through propagating and aggregating needed information. However, it is not trivial for a model to learn hierarchical features that are crucial for graph representation and classification. To address this limitation, we introduced a transformer layer that selects the most significant nodes in the graph and aggregates information via the attention mechanism. Transformers use a Self-Attention (SA) mechanism to model the interactions between all tokens in a sequence [21], by allowing the tokens to interact with each other (“self”) and find out who they should pay more attention to (“attention”), and the addition of positional information of tokens further increases the use of sequential order information. Excitingly, the Vison Transformer (ViT) enables the application of transformers to 2D images [12]. Inspired by these studies, we here propose a transformer layer to interpret our graph-structured data. While the SA mechanism has been extensively used in the context of natural language processing, we extended the framework for WSI data. Briefly, the standard **qkv** self-attention [21] is a mechanism to find the words of importance for a given query word in a sentence, and it receives as input a 1D sequence of token embeddings. For the graph, the feature nodes are treated as tokens in a sequence and the adjacency matrix is used to denote the positional information. Given that **x** ∈ ℝ^*N*×*D*^ is the sequence of patches (feature nodes) in the graph, where *N* is the number of patches and *D* is the embedding dimension of each patch, we compute **q**(query), **k**(key) and **v**(value) (Eq.3a). The attention weights *A*_*ij*_ are based on the pairwise similarity between two patches of the sequence and their respective query **q**^*i*^ and key **k**^*j*^ in Eq.3b. Multihead Self-Attention (MSA) is a mechanism that involves combining the knowledge explored by *k* number of SA operations, called “heads”. It projects concatenated outputs of SA in Eq.3c. *D*_*h*_ (Eq.3a) is typically set to *D/k* to facilitate computation and maintain the number of parameters constant when changing *k*.

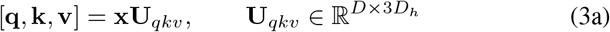

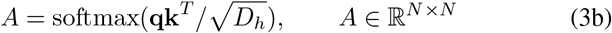

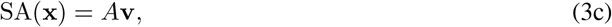

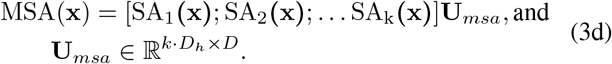

The goal of the transformer layer is to learn the mapping: ℍ →𝕋, where ℍ is the graph space, and T is the transformer space. We define the mapping of ℍ →𝕋 as:

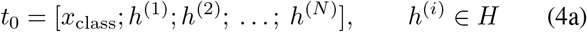

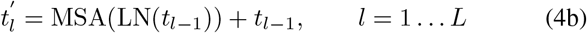

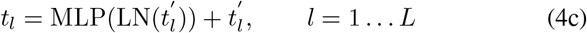

where MSA is the Multiheaded Self-Attention (Eq.3), MLP is a Multilayer Perceptron, and LN denotes Layer Norm. L is the number of MSA blocks [12]. The transformer layer consists of *L* MSA layers (Eq.4b) and *L* MLP blocks (Eq.4c). In order to learn the mapping 𝕋 → 𝕐 from transformer space 𝕋 to label space 𝕐, we prepared a learnable embedding 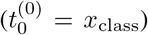 to the feature nodes (Eq.4a), whose state at the output of the transformer layer 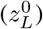 serves as mapping of 𝕋 →𝕐:

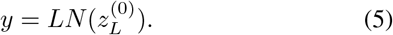

In a recent work [12], position embeddings were added to the patch embeddings to retain positional information. Typically, the position embedding explores absolute position encoding (e.g., sinusoidal encoding, learnable absolute encoding) as well as conditional position encoding. However, the learnable absolute encoding is commonly used in problems with fixed length sequences and does not meet the requirement for variable length of input patches in WSI analysis, because the number of patches tiled from the corresponding WSI often varies due to the inherently variable size of the WSI. To handle this problem, Islam and colleagues showed that the addition of zero padding can provide an absolute position information for convolution [22]. In our work, the adjacency matrix in the WSI graph which contains the spatial information is encoded with the position information and added to the node features during graph convolution. By taking advantage of graph convolutions to aggregate context information, the node features are able to obtain both local and contextual information, which enriches the features that are encompassed in each node. In this fashion, we were able to avoid the need of adding an additional encoder for position embeddings, thus reducing the complexity of our model.

The softmax function is typically used as a row-by-row normalization function in transformers for vision tasks [23], [24]. The standard self-attention mechanism requires the calculation of similarity scores between each pair of nodes, resulting in both memory and time complexity quadratic in the number of nodes. Since the number of patches in WSIs is large (potentially several thousands), applying the transformer layer directly to the convolved graphs is not trivial. We therefore added a mincut pooling layer [25] between the graph convolution and transformer layers and reduced the number of input nodes to the transformer layer. In so doing, our GTP graph-transformer was able to accommodate thousands of image patches as input, which underscores the novelty of our approach and its application to WSI data.

### C. Class activation mapping

To understand how GT processes WSI data and identifies regions that are highly associated with the class label, we proposed a novel class activation mapping technique on graphs. In what follows, we use the term GraphCAM to refer to this technique. Our technique was inspired by the recent work by Chefer and colleagues [26], who used the deep Taylor decomposition principle to assign local relevance scores and propagated them through the layers by maintaining the total relevancy across layers. In a similar fashion, our method computes the class activation map from the output class to the input graph space, and reconstructs the final class activation map for the WSI from its graph representation.

Let *A*^(*l*)^ represent the attention map of the MSA block *l* in Eq.3b. Following the propagation procedure of relevance and gradients by Chefer and colleagues [26], GraphCAM computes the gradient ∇*A*^(*l*)^ and layer relevance 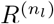 with respect to a target class for each attention map *A*^(*l*)^, where *n*_*l*_ is the layer that corresponds to the softmax operation in Eq.3b of block *l*. The transformer relevance map *C*_*t*_ is then defined as a weighted attention relevance:

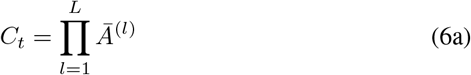

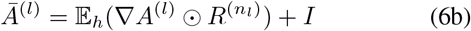

where ⨀ is the Hadamard product, 𝔼_*h*_ is the mean across the “heads” dimension, and *I* is the identity matrix to avoid self inhibition for each node.

The pooled node features by the mincut pooling layer are computed as *X*^*pool*^ = *S*^*T*^ *X*, where 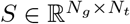 is the dense learned assignment, and *N*_*t*_ and *N*_*g*_ are the number of nodes before and after the pooling layer. To yield the graph relevance map *C*_*g*_ from transformer relevance map *C*_*t*_, our GraphCAM performs mapping *C*_*t*_ to each node in the graph based on the dense learned assignments as 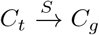. Finally, GraphCAM reconstructs the final class activation map on the input WSI using the adjacency matrix of the graph and coordinates of patches from the WSI.

### D. Data and code availability

All the WSIs and corresponding clinical data can be downloaded freely from CPTAC, TCGA and NLST websites. Python scripts and manuals are made available on GitHub (https://github.com/vkola-lab/GraphCAM).

## III. Experiments

We performed several experiments to train and test our GTP framework. The NLST data (1.8 million patches) was exclusively used for contrastive learning to generate patch-specific features (and the feature extractor), which were then used to represent each node. The GTP framework was trained on the CPTAC data (2, 071 WSIs) using 5-fold cross validation, and the TCGA data (288 WSIs) was used as an independent dataset for model testing using the same hyperparameters. We also conducted ablation studies to understand the contributions of various components on the overall GTP framework. By blocking out the GTP components, we were left with frameworks that were comparable to the state-of-the-art in the field. Finally, we used GraphCAMs to identify salient regions on the WSIs, and explored their validity in terms of highlighting the histopathologic regions of interest.

### A. Experimental settings

Each WSI was cropped to create a bag of 512 × 512 non-overlapping patches at 20 × magnifications, and background patches with non-tissue area *>* 50% were discarded. We used Resnet18 as the CNN backbone used for the feature extractor [27]. We adapted the Adam optimizer with an initial learning rate of 0.0001, a cosine annealing scheme for learning rate scheduling [28], and a mini-batch size of 512. We kept the trained feature extractor and used it to build graphs for the Graph-Transformer. We used one graph convolutional layer, and set the transformer layer configurations as *L*=3, MLP *size*=128, *D*=64 and *k*=8 (Eq.4, Eq.3). The GTP model was trained in batches of 8 examples for 150 iterations. We adopted Adam [29] as the optimizer. The learning rate was set to 10^− 3^ initially, and decayed to 10^− 4^ and 10^− 5^ at step 30 and 100, respectively.

### B. Ablation studies

We compared the effect of contrastive learning on the GTP model performance by performing studies with and without it. Later, we removed the transformer component and trained the graph and compared it with the full GTP framework. In both these studies, we explored various options to build the model, including the use of pre-training to generate the features in lieu of contrastive learning, and also used a graph-based CNN to predict the class label as a replacement to the transformer. In essence, these ablation studies allowed us to fully evaluate the power of our interpretable GTP framework in predicting WSI-level class labels.

### C. Computational infrastructure

We implemented the proposed model using PyTorch (v1.9.0). The model was trained using a single NVIDIA 1080Ti graphics card with 12 GB memory on a GPU workstation. The training speed was about 2.4 iterations/s, and training took less than a day to reach convergence. The inference speed was about 30 ms per WSI when the test batch size was 2.

### D. Performance metrics

For the tumor versus normal classification task, we generated receiver operating characteristic (ROC) and precision-recall (PR) curves based on model predictions on the CPTAC and TCGA datasets. The ROC curve was computed between the true positive rate and false positive rate using different probability thresholds while PR curve was computed between the true positive rate and the positive predictive value using different probability thresholds. For each ROC and PR curve, we also computed the area under curve (AUC), precision, recall, specificity, and accuracy. For the 3-label classification task (LUAD vs. LSCC vs. normal), we also computed the precision, recall, specificity, and accuracy scores of each class along with confusion matrices for each fold-level prediction. The ROC and PR curves were computed for each label. Since we used 5-fold cross validation, we took all the curves from different folds and calculated the mean area under curves and the variance of the curves. Finally, GraphCAMs were used to generate visualizations and gain a qualitative understanding on the model performance.

## IV. Results

The GTP framework that leveraged contrastive learning followed by fusion of a graph with a transformer provided accurate predictions of WSI-level class labels across a range of classification tasks (Table II). For the normal vs. tumor classification task, high model performance was consistently observed on all the computed metrics including precision, recall, sensitivity, and overall accuracy on both CPTAC test and TCGA datasets (all > 0.9), indicating a high degree of generalizability. Similar performance was observed on the normal vs. LUAD vs. LSCC task on the CPTAC data but dropped slightly on the TCGA dataset. The drop in the model performance was observed particularly on the precision scores for the LUAD and on the recall scores for the LSCC class labels. High model performance was also confirmed via the receiver-operating characteristic (ROC) and precision-recall (PR) curves generated on both the CPTAC and TCGA datasets for all the classification tasks (Figure 3). On each classification task, the mean area under the ROC and PR curves was high (all > 0.9) on the CPTAC test data. For the TCGA dataset, which served as an external testing cohort, the mean area under the ROC and PR curves dropped slightly, especially for the LUAD and LSCC classification tasks. The confusion matrices for the 3-label classification problem indicated similar results (Figure S3), where the model performance was excellent on the CPTAC test dataset but was slightly lower on the TCGA dataset. In particular, the model leaned towards incorrectly classifying a few LSCC cases as LUAD but correctly classified most of the WSIs with no tumor. However, for the two-label classification task (i.e., tumor vs. no-tumor), the mean area under the ROC and PR curves were very high on both the CPTAC and TCGA datasets (all > 0.95), indicating accurate model performance and a fair degree of model generalizability (Figure S4).

**TABLE II:** Performance metrics for each class in the 3-label and 2-label classification tasks.

(a) 2-label: Normal vs. Tumor (LUAD+LSCC)
| CPTAC | Precision | Recall/Sensitivity | Specificity |
| --- | --- | --- | --- |
| Normal | $0.953 \pm 0.022$ | $0.978 \pm 0.017$ | $0.974 \pm 0.012$ |
| Tumor | $0.988 \pm 0.009$ | $0.974 \pm 0.012$ | $0.978 \pm 0.017$ |
| TCGA | Precision | Recall/Sensitivity | Specificity |
| Normal | $0.902 \pm 0.020$ | $0.937 \pm 0.023$ | $0.952 \pm 0.011$ |
| Tumor | $0.970 \pm 0.011$ | $0.952 \pm 0.011$ | $0.937 \pm 0.023$ |

(b) 3-label: Normal vs. LUAD vs. LSCC
| CPTAC | Precision | Recall/Sensitivity | Specificity |
| --- | --- | --- | --- |
| Normal | $0.953 \pm 0.022$ | $0.978 \pm 0.017$ | $0.974 \pm 0.012$ |
| LUAD | $0.925 \pm 0.032$ | $0.901 \pm 0.021$ | $0.965 \pm 0.016$ |
| LSCC | $0.919 \pm 0.022$ | $0.915 \pm 0.042$ | $0.959 \pm 0.011$ |
| TCGA | Precision | Recall/Sensitivity | Specificity |
| Normal | $0.902 \pm 0.020$ | $0.937 \pm 0.023$ | $0.952 \pm 0.011$ |
| LUAD | $0.747 \pm 0.022$ | $0.841 \pm 0.032$ | $0.854 \pm 0.021$ |
| LSCC | $0.885 \pm 0.015$ | $0.741 \pm 0.031$ | $0.949 \pm 0.009$ |

(c) Overall accuracy
| CPTAC | Accuracy | TCGA | Accuracy |
| --- | --- | --- | --- |
| 2-label | $0.975 \pm 0.013$ | 2-label | $0.947 \pm 0.011$ |
| 3-label | $0.932 \pm 0.019$ | 3-label | $0.838 \pm 0.009$ |

**Fig. 2:**
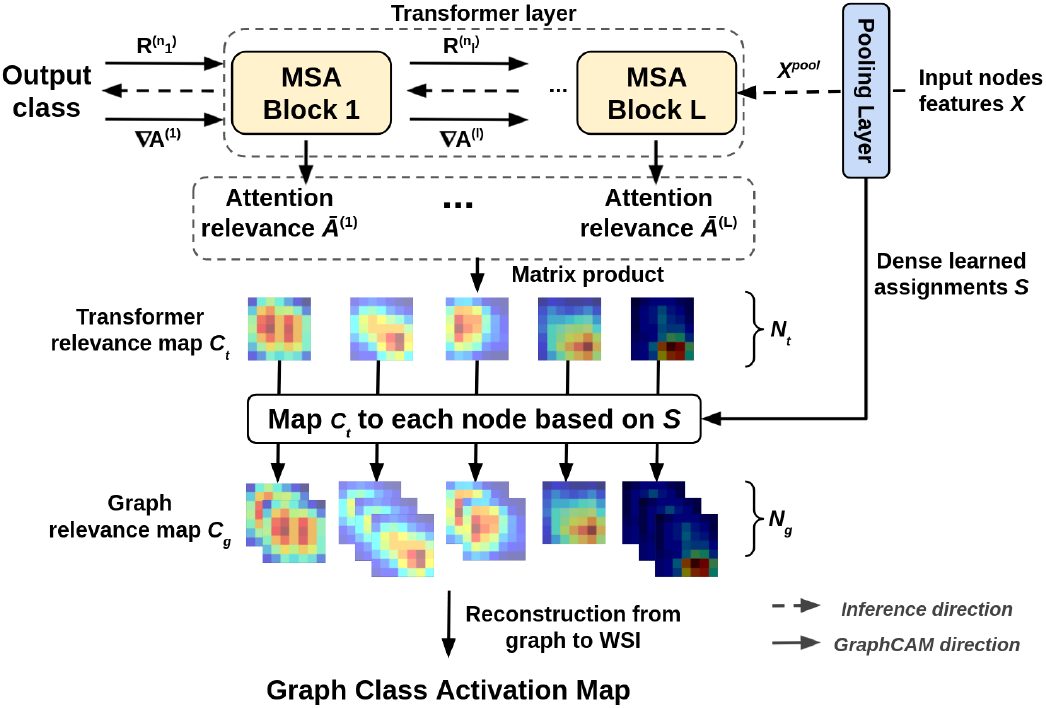
Schematic of the GraphCAM. Gradients and relevance are propagated through the network and integrated with an attention map to produce the transformer relevancy maps. Transformer relevancy maps are then mapped to graph class activation maps via reverse pooling.

**Fig. 3:**
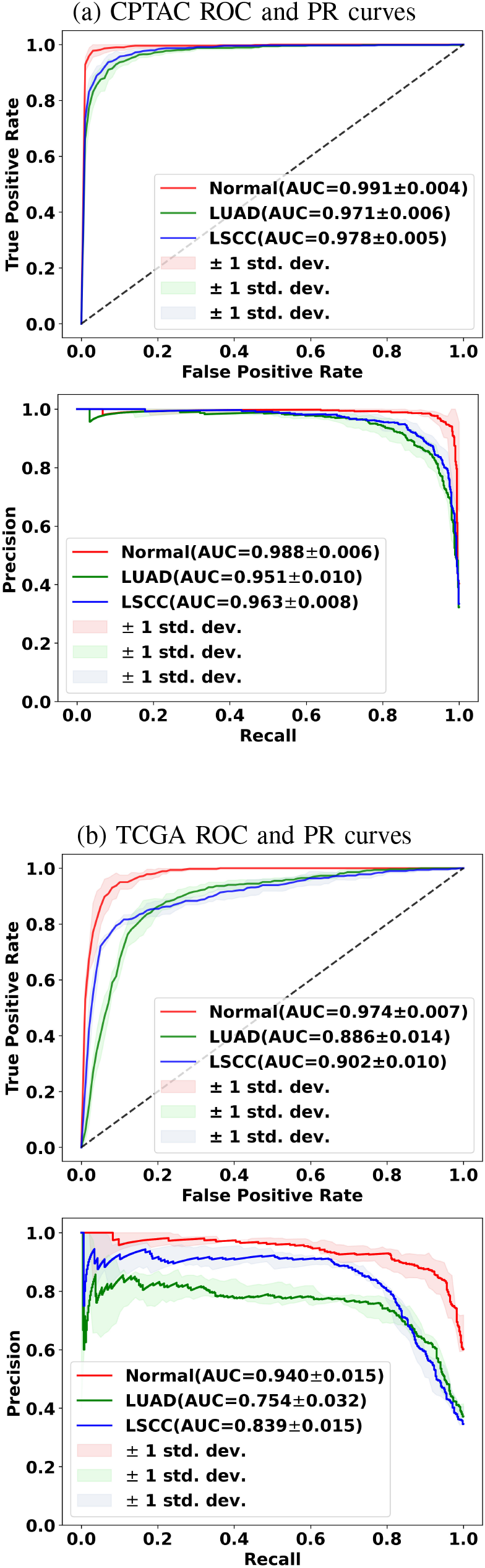
Model performance on the (a) CPTAC and (b) TCGA datasets. Mean ROC and PR curves along with standard deviations for the classification tasks (normal vs. tumor; LUAD vs. others; LSCC vs. others) are shown.

The GT-based class activation maps (GraphCAMs) identified WSI regions that were highly associated with the output class label of interest (Figure 4). Importantly, the same set of WSI regions were highlighted by our method across the various cross-validation folds (Figure S5), thus indicating consistency of our technique in highlighting salient regions of interest. Also, the generated GraphCAMs are class-specific, thus underscoring the superiority of our technique compared to other state-of-the-art methods such as self-attention maps. In some cases, we also noticed that the GraphCAMs generated for each class identified different regions of importance on the same WSI, raising the possibility that a single image may contain disease related information relevant to multiple types of lung cancer. On the other hand, the self-attention map that combines attention across all the layers of the model resulted in a single heatmap that may not indicate disease specificity but rather only an association with the classification task. Also, since we can generate class-specific probability for each GraphCAM, our approach allows for better appreciation of the model performance and its interpretability in predicting an output class label. We must however note that in certain cases when the model fails to predict the class label, the GraphCAMs may not result in interpretable findings (Figure S6). Nevertheless, we note that the saliency maps reported in Figure 4 closely match with expert-identified regions of tumor pathology.

**Fig. 4:**
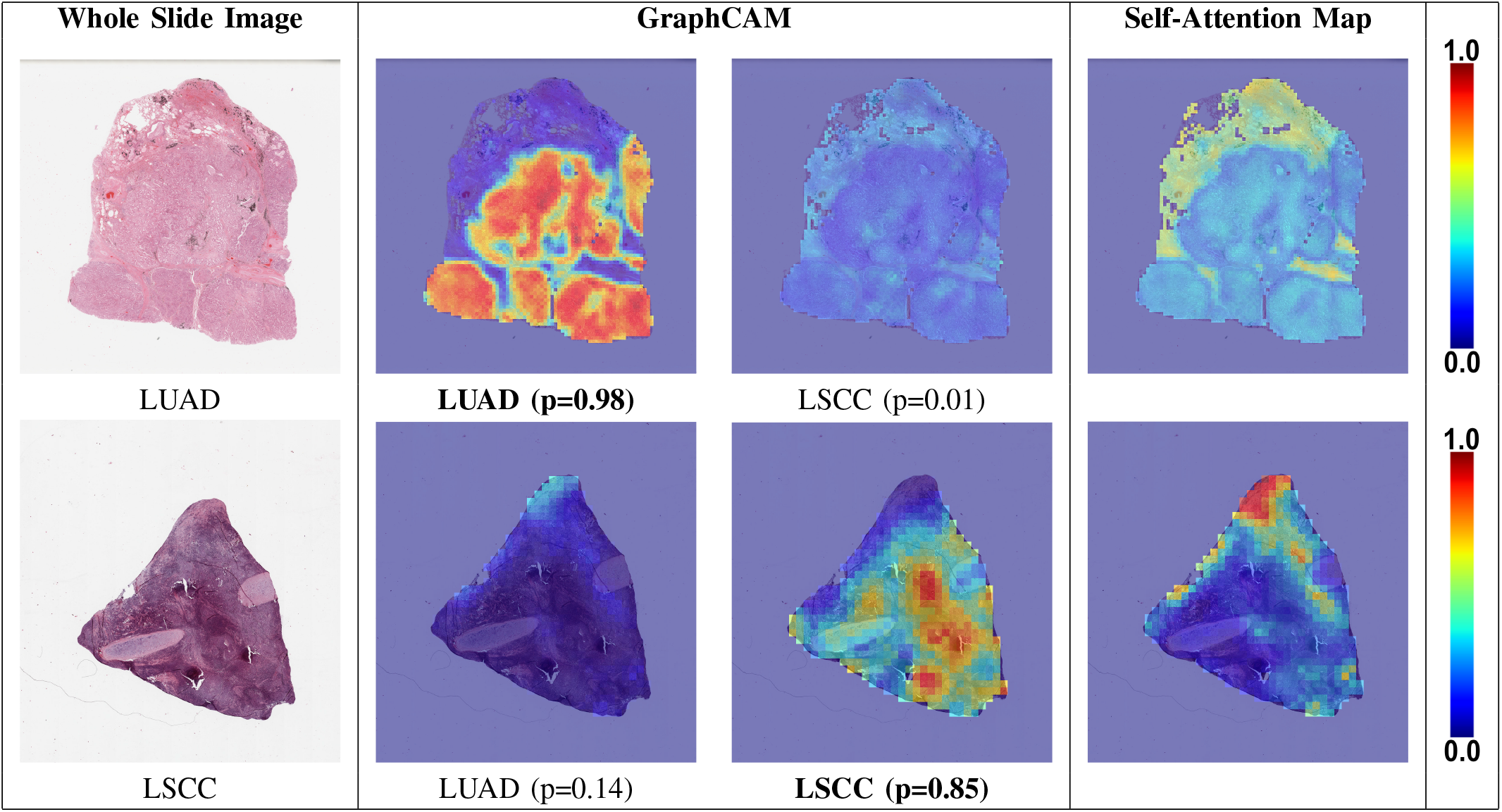
Class-specific GraphCAMs. For each WSI, we generated class-specific GraphCAMs and also compared them with self-attention maps. The first column contains the original WSIs, the second and third columns contain LUAD-specific and LSCC-specific GraphCAMs, and the final column contains the self-attention maps. The first row represents an LUAD case where our model also predicted LUAD, and the second row represents an LSCC case where our model predicted LSCC. The bold font underneath certain GraphCAMs was used to indicate the model predicted class label for the respective cases. Also, the model-generated probability values are noted beneath each GraphCAM. Since this is a 3-label classification task (normal vs. LUAD vs. LSCC), the LUAD and LSCC probability values do not add up to 1. Several patches in the GraphCAM with the correct prediction show high values (“warm colors”) while the self attention maps mostly miss the relevant patches (except in the last row).

Ablation studies revealed that our GTP framework that uses contrastive learning and combines a graph with a transformer served as a superior model for WSI-level classification (Table III). For example, when contrastive learning was replaced with a pre-trained architecture (Resnet18 with and without fine tuning), the model performance for both the 2- and 3-label classification tasks dropped. The reduction in performance was evident on both CPTAC and TCGA datasets. The model performance also dropped for both 2- and 3-label classification when we trained a novel convolutional auto-encoder [30] in lieu of contrastive learning. These results imply that the feature maps generated via contrastive learning were sufficient and maybe even better than other frameworks to encode a large variety of visual information for GT-based classification with a sufficient degree of generalizability. We also replaced our proposed mincut pooling with an attention-based pooling (AttPool) layer that selects the most significant nodes in the graph and aggregates information via the attention mechanism. We then used the same graph convolutional layer as GTP in the ablation study and denoted this method as GraphAtt. By aggregating the neighborhood node information via selfattention, GTP outperformed GraphAtt for both the 2- and 3-label classification tasks (Table S1). These findings indicate that our proposed GTP framework is capable of integrating information across the entire WSI that is represented as a graph to accurately predict the output label of interest.

**TABLE III:** Ablation studies. We used different feature extractors for graph construction also explored the effect of using the transformer by replacing it with a graph classifier. Here, Resnet^*^ indicates the use of a pre-trained Resnet18 network without fine-tuning. Also, Resnet† indicates the use of a pre-trained Resnet18 with fine-tuning. CAE represents convolutional auto encoder, CL represents contrastive learning used in our method and GT represents the Graph-Transformer. Overall values of mean accuracy ± standard deviation, computed across the five folds, are computed.

| CPTAC | Label | 2-label |  |  | Label | 3-label |  |  |
| --- | --- | --- | --- | --- | --- | --- | --- | --- |
|  |  | Precision | Recall/Sensitivity | Specificity |  | Precision | Recall/Sensitivity | Specificity |
| Resnet<sup>*</sup> + GT | Normal | 0.874 ± 0.051 | 0.912 ± 0.029 | 0.927 ± 0.035 | Normal | 0.874 ± 0.051 | 0.912 ± 0.029 | 0.927 ± 0.035 |
| | Tumor | 0.953 ± 0.013 | 0.927 ± 0.035 | 0.912 ± 0.029 | LUAD | 0.809 ± 0.082 | 0.725 ± 0.105 | 0.910 ± 0.047 |
| | | | | | LSCC | 0.786 ± 0.057 | 0.800 ± 0.090 | 0.885 ± 0.055 |
| Resnet<sup>†</sup> + GT | Normal | 0.901 ± 0.032 | 0.915 ± 0.031 | 0.946 ± 0.019 | Normal | 0.901 ± 0.032 | 0.915 ± 0.031 | 0.946 ± 0.019 |
| | Tumor | 0.955 ± 0.015 | 0.946 ± 0.019 | 0.915 ± 0.031 | LUAD | 0.771 ± 0.030 | 0.756 ± 0.035 | 0.894 ± 0.013 |
| | | | | | LSCC | 0.790 ± 0.039 | 0.788 ± 0.022 | 0.894 ± 0.028 |
| CAE + GT | Normal | 0.893 ± 0.017 | 0.864 ± 0.042 | 0.944 ± 0.012 | Normal | 0.893 ± 0.017 | 0.864 ± 0.042 | 0.944 ± 0.012 |
| | Tumor | 0.930 ± 0.020 | 0.944 ± 0.012 | 0.864 ± 0.042 | LUAD | 0.733 ± 0.041 | 0.674 ± 0.058 | 0.881 ± 0.031 |
| | | | | | LSCC | 0.753 ± 0.028 | 0.829 ± 0.036 | 0.863 ± 0.027 |
| CL + GraphAtt | Normal | 0.878 ± 0.024 | 0.902 ± 0.028 | 0.933 ± 0.015 | Normal | 0.878 ± 0.024 | 0.902 ± 0.028 | 0.933 ± 0.015 |
| | Tumor | 0.948 ± 0.014 | 0.933 ± 0.015 | 0.902 ± 0.028 | LUAD | 0.791 ± 0.027 | 0.758 ± 0.063 | 0.905 ± 0.017 |
| | | | | | LSCC | 0.835 ± 0.046 | 0.841 ± 0.032 | 0.916 ± 0.029 |
| TCGA | Label | 2-label |  |  | Label | 3-label |  |  |
|  |  | Precision | Recall/Sensitivity | Specificity |  | Precision | Recall/Sensitivity | Specificity |
| Resnet<sup>*</sup> + GT | Normal | 0.639 ± 0.203 | 0.146 ± 0.116 | 0.964 ± 0.037 | Normal | 0.639 ± 0.203 | 0.146 ± 0.116 | 0.964 ± 0.037 |
| | Tumor | 0.707 ± 0.027 | 0.964 ± 0.037 | 0.146 ± 0.116 | LUAD | 0.375 ± 0.009 | 0.893 ± 0.052 | 0.244 ± 0.050 |
| | | | | | LSCC | 0.695 ± 0.098 | 0.257 ± 0.040 | 0.941 ± 0.019 |
| Resnet<sup>†</sup> + GT | Normal | 0.638 ± 0.036 | 0.765 ± 0.022 | 0.794 ± 0.034 | Normal | 0.638 ± 0.036 | 0.765 ± 0.022 | 0.794 ± 0.034 |
| | Tumor | 0.878 ± 0.010 | 0.794 ± 0.034 | 0.765 ± 0.022 | LUAD | 0.515 ± 0.019 | 0.536 ± 0.039 | 0.742 ± 0.033 |
| | | | | | LSCC | 0.662 ± 0.024 | 0.509 ± 0.060 | 0.865 ± 0.014 |
| CAE + GT | Normal | 0.620 ± 0.019 | 0.894 ± 0.027 | 0.742 ± 0.025 | Normal | 0.620 ± 0.019 | 0.894 ± 0.027 | 0.742 ± 0.025 |
| | Tumor | 0.937 ± 0.014 | 0.742 ± 0.025 | 0.894 ± 0.027 | LUAD | 0.501 ± 0.062 | 0.342 ± 0.067 | 0.821 ± 0.051 |
| | | | | | LSCC | 0.598 ± 0.014 | 0.529 ± 0.074 | 0.813 ± 0.034 |
| CL + GraphAtt | Normal | 0.826 ± 0.016 | 0.913 ± 0.017 | 0.909 ± 0.008 | Normal | 0.826 ± 0.016 | 0.913 ± 0.017 | 0.909 ± 0.008 |
| | Tumor | 0.957 ± 0.008 | 0.909 ± 0.008 | 0.913 ± 0.017 | LUAD | 0.718 ± 0.040 | 0.753 ± 0.035 | 0.850 ± 0.023 |
| | | | | | LSCC | 0.858 ± 0.027 | 0.732 ± 0.026 | 0.937 ± 0.012 |

(b) Accuracy
|  |  | Resnet<sup>*</sup> + GT | Resnet<sup>†</sup> + GT | CAE + GT | CL + GraphAtt |
| --- | --- | --- | --- | --- | --- |
| CPTAC | 2-label | 0.922 ± 0.017 | 0.935 ± 0.010 | 0.917 ± 0.009 | 0.923 ± 0.013 |
| | 3-label | 0.815 ± 0.010 | 0.822 ± 0.019 | 0.791 ± 0.011 | 0.835 ± 0.022 |
| TCGA | 2-label | 0.703 ± 0.036 | 0.785 ± 0.022 | 0.790 ± 0.015 | 0.911 ± 0.011 |
| | 3-label | 0.435 ± 0.022 | 0.600 ± 0.021 | 0.583 ± 0.018 | 0.797 ± 0.026 |

## V. Discussion

In this work, we developed a novel deep learning approach that integrates graphs with vision transformers to generate an efficient classifier to predict WSI-level presence of lung tumors. Our approach also differentiated WSIs with LUAD from those with LSCC. Based on the standards of various model performance metrics, our approach resulted in classification performance that exceeded other deep learning architectures that incorporated various state-of-the-art configurations (see ablation studies). Finally, our novel class activation mapping technique allowed us to identify salient WSI regions that were highly associated with the output class label of interest. Thus, our findings represent novel contributions to the field of in-terpretable deep learning while also simultaneously advancing the fields of computer vision and digital pathology.

The field of computational pathology has made important strides in the recent years due to advancements in vision-based deep learning. Still, owing to the sheer size of pathology images generated at high resolution, assessment of WSI-level information that can integrate spatial signatures along with local, region-specific information for prediction of tumor grade remains a challenge. The large body of work to date has focused on patch-level deep neural networks that may accurately predict tumor grade but fail to capture spatial connectivity information. As a result, identification of important image-level features via such techniques may lead to inconsistent results. Our GT-based deep learning framework precisely tackled this scenario by integrating WSI-level information via a graph structure and thus represents an important advancement in the field.

One of the novel contributions in our work is the generation of graph-based class activation maps (GraphCAM), which can highlight the WSI regions that are highly associated with the output class label. Unlike other saliency mapping techniques such as self-attention maps, GraphCAMs can generate class-specific heatmaps. While self-attention maps can identify image regions (or pixels) that are important for a specific classification task, GraphCAMs can identify image regions that trigger the model to associate the image with a specific class label. This is a major advantage because an image may contain information pertaining to multiple classes, and for these scenarios, identification of class-specific feature maps becomes important. This is especially true in real-world scenarios such as pathology images containing lung tumors. Typically, lung cancer subtype on WSIs is determined based on the most predominant pattern, but different patterns may be present on the same WSIs. In such cases, training well-known supervised deep learning classifiers such as convolutional neural networks that use the overall WSI label for classification at patch-level or even at the WSI-level may not necessarily perform well and even misidentify the regions of interest associated with the class label. By generating class-specific CAMs learned at the WSI-level, our GTP approach provides an accurate way by which to identify regions of interest on WSIs that are highly associated with the corresponding class label.

Our study has a few limitations. We leveraged contrastive learning to generate patch-level feature vectors before constructing the graph, which turned out to be a computationally intensive task. However, our ablation studies revealed that contrastive learning improved model performance when compared to other techniques for feature extraction. Future studies can explore other possible techniques for feature extraction that lead to improved model performance. Our graph was constructed by dividing the WSI into image patches, followed by creation of nodes using the embedding features from these patches leading to construction of the graph. Other alternative ways can be explored to define the nodes and create graphs that are more congruent and spatially connected. While we have demonstrated the applicability of GTP to lung tumors, extension of this framework to other cancers is needed to fully appreciate its role in terms of assessing WSI-level correlates of disease. In fact, our method is not specific to cancers and could be adapted to other computational pathology applications.

In conclusion, our GTP framework produced an accurate, computationally efficient model by capturing the entire information available on an WSI to predict the output class label. As a supervised learning framework, GTP can tackle large resolution WSIs and predict multiple class labels, leading to generation of interpretable findings that are class-specific. Our GTP framework could be scaled to WSI-level classification tasks on other organ systems and also to predict response to therapy, cancer recurrence and patient survival.

## Data Availability

TCGA, NLST and CPTAC websites

## Supplement

**Fig. S1:**
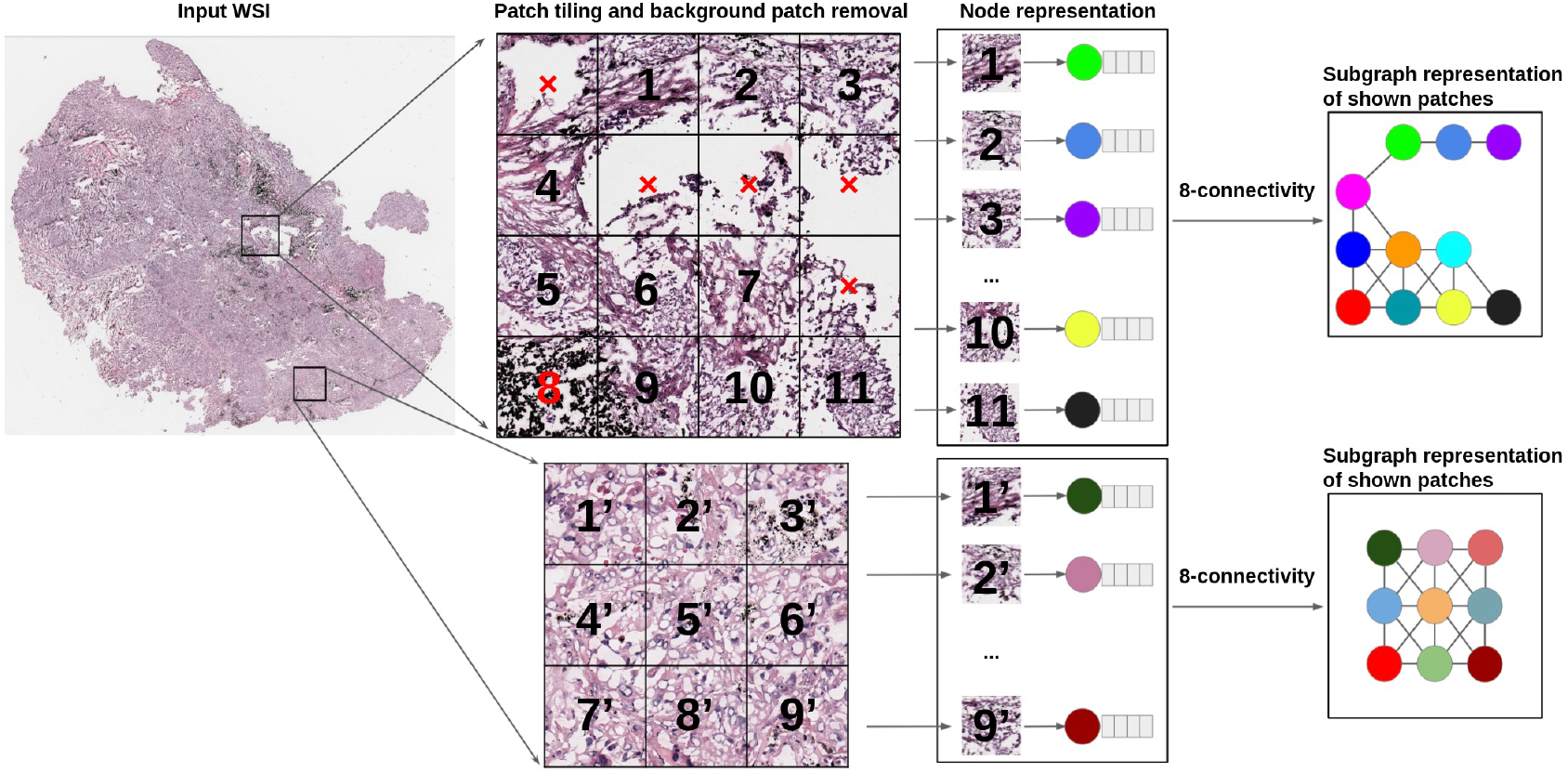
Graph construction. Whole slide images were divided into patches and each patch that contained more than 50% of the area covered by tissue was considered for further processing. Each selected patch was represented as a node and a graph was constructed on the entire WSI using these nodes with an 8-node adjacency matrix. Here, two sets of patches of a WSI and their corresponding subgraphs are shown. The subgraphs are connected within the graph representing the entire WSI.

**Fig. S2:**
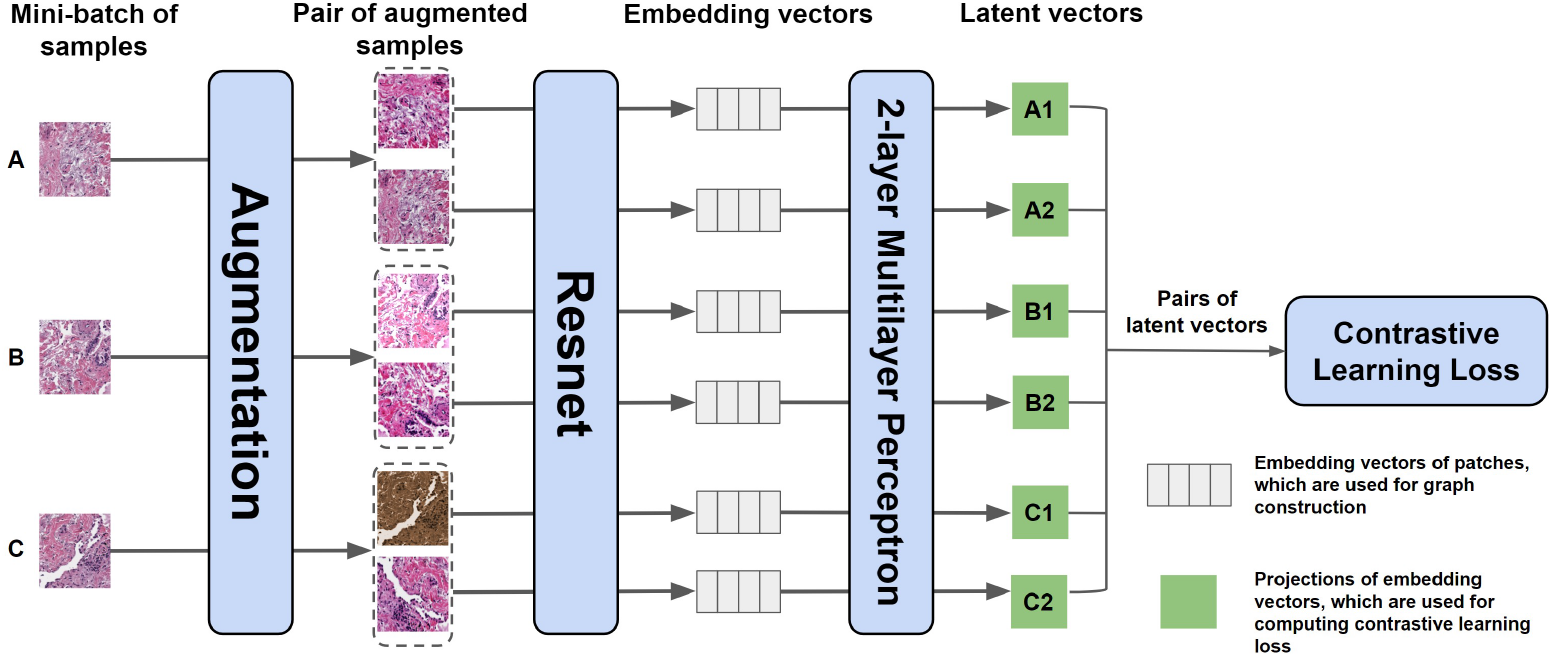
Contrastive learning to train the feature extractor. We applied two distinct augmentation functions, including random color distortions, random Gaussian blur, and random cropping followed by resizing back to the original size, on the same sample in a mini-batch. If the mini-batch size is *K*, then we ended up with 2 × *K* augmented observations in the mini-batch. The ResNet received an augmented image leading to an embedding vector as the output. Subsequently, a projection head was applied to the embedding vector which produced the inputs to contrastive learning. The projection head is a multilayer perceptron (MLP) with 2 dense layers. In this example, we considered *K* = 3 samples in a minibatch (A, B & C). For the sample A, the positive pairs are (A1, A2) and (A2, A1), and the negative pairs are (A1, B1), (A1, B2), (A1, C1), (A1, C2). All pairs were used for computing contrastive learning loss to train the Resnet. Once the system was trained, we used the embedding vectors (straight from the ResNet) for constructing the graph.

**Fig. S3:**
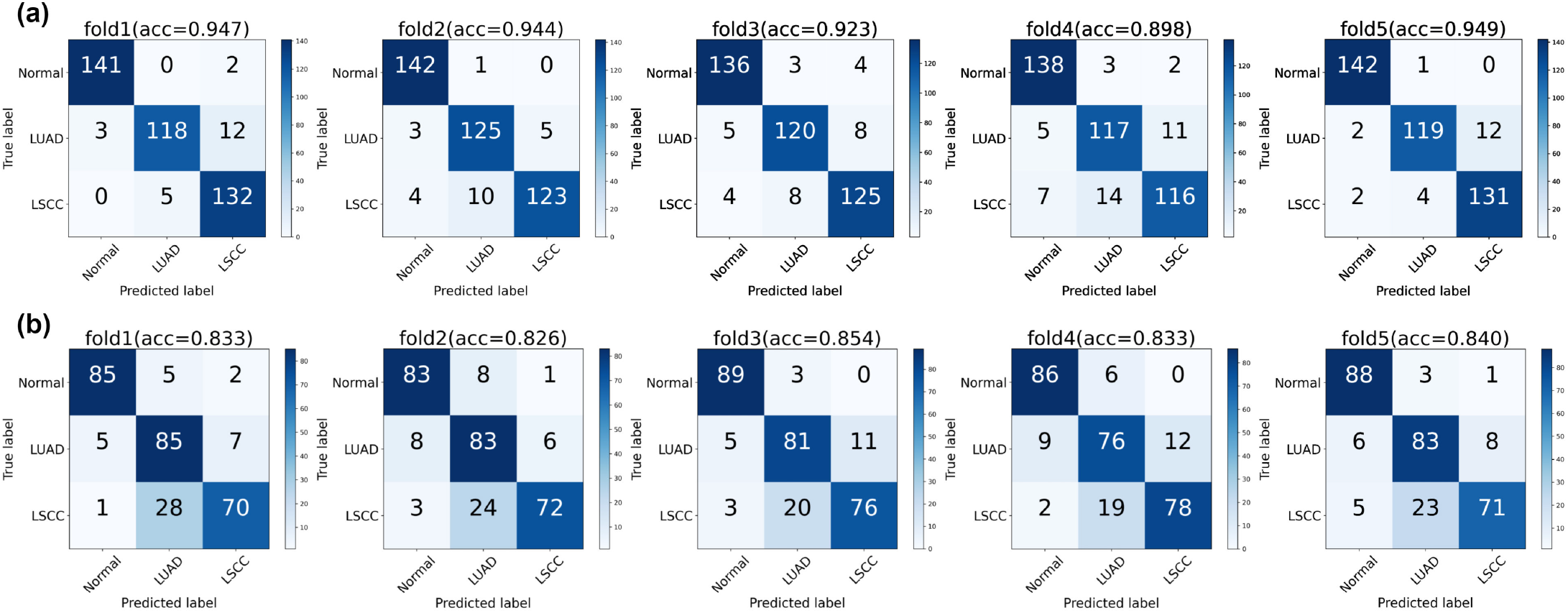
Model performance on the CPTAC and TCGA datasets. Confusion matrices for the 5-fold cross validation on the (a) CPTAC and the (b) TCGA datasets are shown. A separate confusion matrix is shown for each fold prediction along with corresponding accuracies. Note that for CPTAC dataset, the model performance was evaluated only on the held-out test data. Model performance on the TCGA dataset was evaluated using the CPTAC model that was constructed on each fold.

**Fig. S4:**
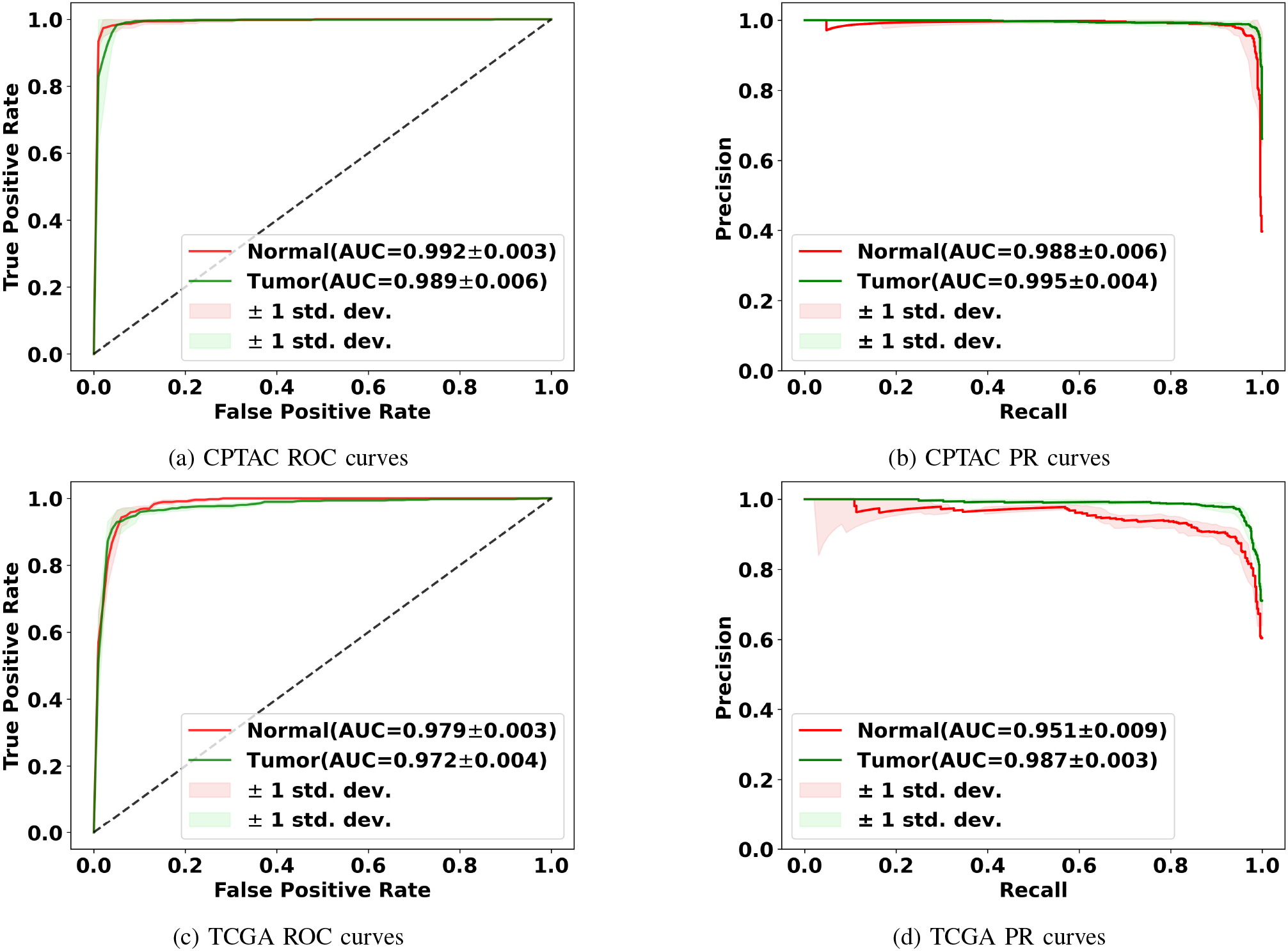
Model performance on the CPTAC and TCGA dataset. Mean ROC and PR curves along with standard deviations for the binary classification task (normal vs. tumor) are shown.

**Fig. S5:**
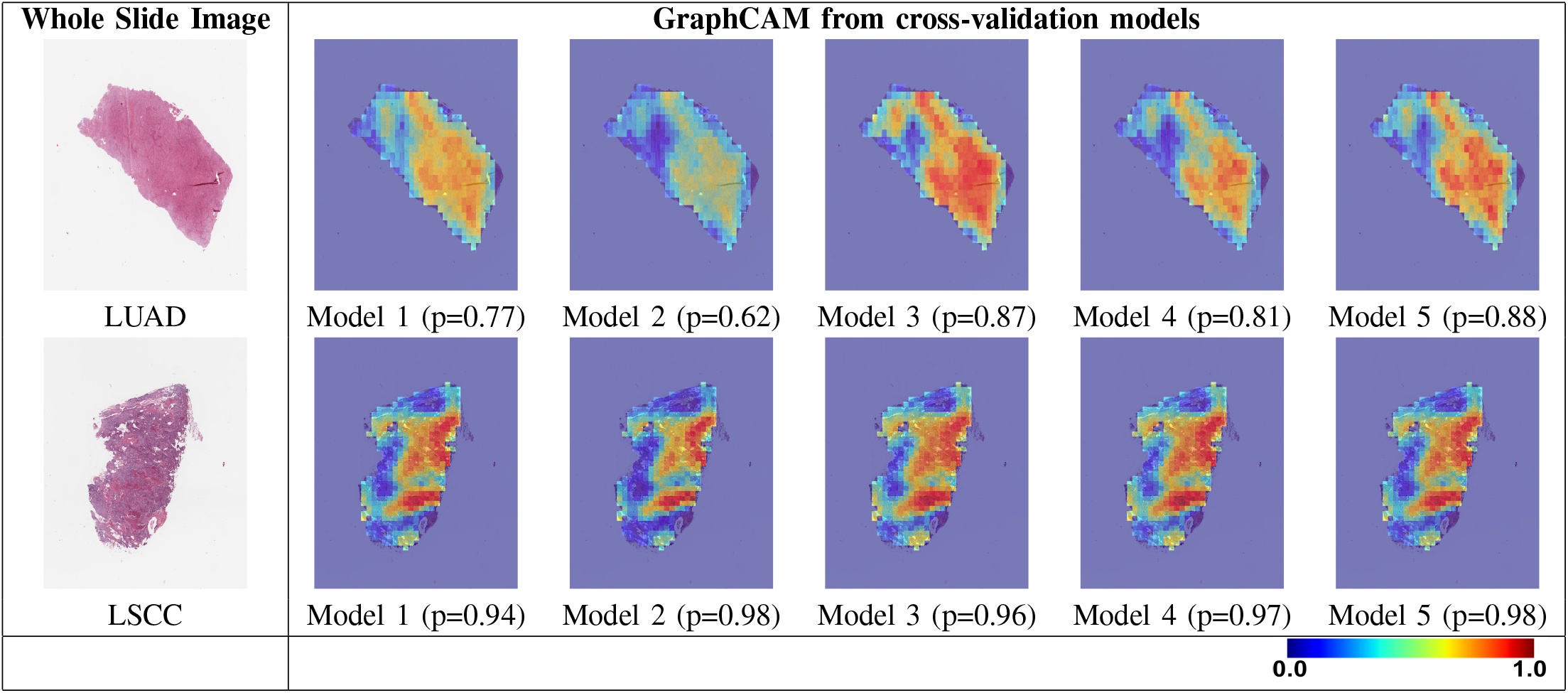
Graph class activation map (GraphCAM) performance. GraphCAMs generated on WSIs across the runs performed via 5-fold cross validation are shown. The first column shows the original WSI and the other columns show the generated GraphCAMs along with prediction probabilities on the cross-validated model runs. The first row shows a sample WSI from the LUAD class and the second row shows an WSI from the LSCC class. The colormap of the GraphCAM represents the probability by which an WSI region is associated with the output label of interest. The probability values based on each model prediction are noted beneath each GraphCAM. The models created with the five folds produced GraphCAMs that mostly highlighted the same set of patches at similar levels, thus underscoring the robustness of our method across the folds.

**Fig. S6:**
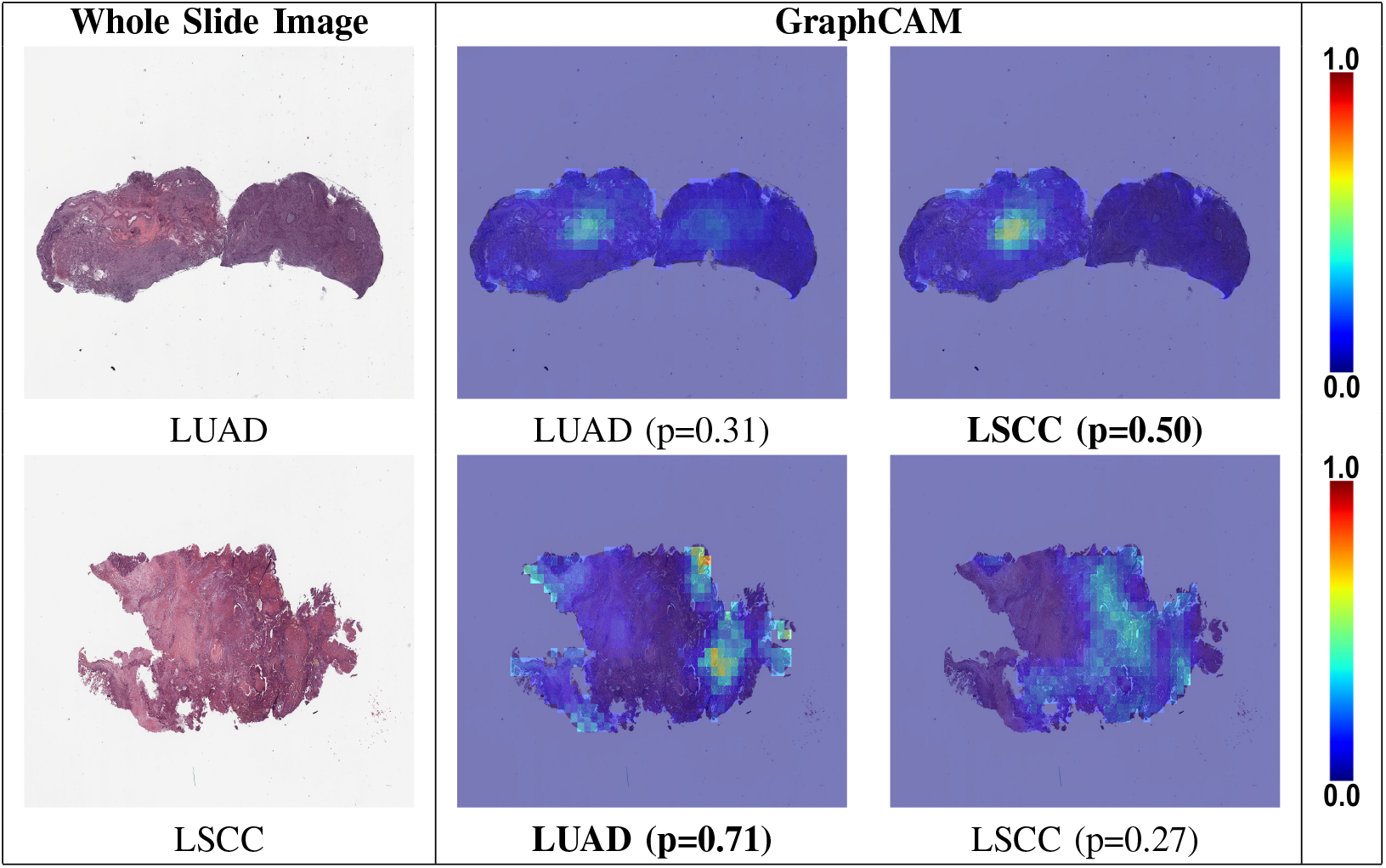
Class-specific GraphCAMs for failure cases. The first row shows a sample WSI from the LUAD class but the model prediction was LSCC, and the second row shows an WSI from the LSCC class but the model prediction was LUAD. The first column shows the original WSI, and the second and third columns show the generated GraphCAMs along with prediction probabilities. The bold font underneath certain GraphCAMs was used to indicate the model predicted class label for the respective cases. Also, the model-generated probability values are noted beneath each GraphCAM. Since this is a 3-label classification task (normal vs. LUAD vs. LSCC), the LUAD and LSCC probability values do not add up to 1.

**TABLE S1:** Two-tailed DeLong test to compare AUCs. We used the DeLong test to compare the AUC values of the models used in the ablation studies. For the ablation studies, we used different feature extractors for graph construction also explored the effect of using the transformer by replacing it with a graph classifier. Here, Resnet^*^ indicates the use of a pre-trained Resnet18 network without fine-tuning. Also, Resnet^†^ indicates the use of a pre-trained Resnet18 with fine-tuning. CAE represents convolutional auto encoder, CL represents contrastive learning used in our method and GT represents the Graph-Transformer. Overall values of mean z-statistic ± standard deviation obtained from the DeLong test are reported. The symbol ^‡^ indicates p-value < 0.001, and § indicates p-value < 0.005. When the CL + GraphAtt model was compared with our model, the p-value was 0.209 on the TCGA dataset.

|  | Resnet<sup>*</sup> + GT | Resnet<sup>†</sup> + GT | CAE + GT | CL + GraphAtt |
| --- | --- | --- | --- | --- |
| CPTAC | $3.208 \pm 0.378^{\ddagger}$ | $2.995 \pm 0.633^{\S}$ | $3.524 \pm 0.561^{\ddagger}$ | $3.503 \pm 0.607^{\ddagger}$ |
| TCGA | $10.949 \pm 3.742^{\ddagger}$ | $5.383 \pm 0.373^{\ddagger}$ | $5.622 \pm 0.239^{\ddagger}$ | $1.256 \pm 0.492$ |

